# Heterozygous loss-of-function *SMC3* variants are associated with variable and incompletely penetrant growth and developmental features

**DOI:** 10.1101/2023.09.27.23294269

**Authors:** Morad Ansari, Kamli N. W. Faour, Akiko Shimamura, Graeme Grimes, Emeline M. Kao, Erica R. Denhoff, Ana Blatnik, Daniel Ben-Isvy, Lily Wang, Benjamin M. Helm, Helen Firth, Amy M. Breman, Emilia K. Bijlsma, Aiko Iwata-Otsubo, Thomy J.L. de Ravel, Vincent Fusaro, Alan Fryer, Keith Nykamp, Lara G. Stühn, Tobias B. Haack, G. Christoph Korenke, Panayiotis Constantinou, Kinga M. Bujakowska, Karen J. Low, Emily Place, Jennifer Humberson, Melanie P. Napier, Jessica Hoffman, Jane Juusola, Matthew A. Deardorff, Wanqing Shao, Shira Rockowitz, Ian Krantz, Maninder Kaur, Sarah Raible, Sabine Kliesch, Moriel Singer-Berk, Emily Groopman, Stephanie DiTroia, Sonia Ballal, Siddharth Srivastava, Kathrin Rothfelder, Saskia Biskup, Jessica Rzasa, Jennifer Kerkhof, Haley McConkey, Anne O’Donnell-Luria, Bekim Sadikovic, Sarah Hilton, Siddharth Banka, Frank Tüttelmann, Donald Conrad, Michael E. Talkowski, David R. FitzPatrick, Philip M. Boone

## Abstract

Heterozygous missense variants and in-frame indels in *SMC3* are a cause of Cornelia de Lange syndrome (CdLS), marked by intellectual disability, growth deficiency, and dysmorphism, via an apparent dominant-negative mechanism. However, the spectrum of manifestations associated with *SMC3* loss-of-function variants has not been reported, leading to hypotheses of alternative phenotypes or even developmental lethality. We used matchmaking servers, patient registries, and other resources to identify individuals with heterozygous, predicted loss-of-function (pLoF) variants in *SMC3*, and analyzed population databases to characterize mutational intolerance in this gene. Here, we show that *SMC3* behaves as an archetypal haploinsufficient gene: it is highly constrained against pLoF variants, strongly depleted for missense variants, and pLoF variants are associated with a range of developmental phenotypes. Among 13 individuals with *SMC3* pLoF variants, phenotypes were variable but coalesced on low growth parameters, developmental delay/intellectual disability, and dysmorphism reminiscent of atypical CdLS. Comparisons to individuals with *SMC3* missense/in-frame indel variants demonstrated a milder presentation in pLoF carriers. Furthermore, several individuals harboring pLoF variants in *SMC3* were nonpenetrant for growth, developmental, and/or dysmorphic features, some instead having intriguing symptomatologies with rational biological links to *SMC3* including bone marrow failure, acute myeloid leukemia, and Coats retinal vasculopathy. Analyses of transcriptomic and epigenetic data suggest that *SMC3* pLoF variants reduce *SMC3* expression but do not result in a blood DNA methylation signature clustering with that of CdLS, and that the global transcriptional signature of *SMC3* loss is model-dependent. Our finding of substantial population-scale LoF intolerance in concert with variable penetrance in subjects with *SMC3* pLoF variants expands the scope of cohesinopathies, informs on their allelic architecture, and suggests the existence of additional clearly LoF-constrained genes whose disease links will be confirmed only by multi-layered genomic data paired with careful phenotyping.

## Introduction

The cohesin complex, a multimeric structure with the ability to entrap DNA, is integral to several dynamic genome processes.^1,2^ Specifically, cohesin facilitates sister chromatid cohesion during cell division,^3^ DNA repair,^4^ 3D chromatin architecture,^5^ and transcriptional control of developmental genes.^6,7^ SMC3 (structural maintenance of chromosomes 3), the sole subunit shared by mitotic, interphase, and meiotic cohesin, is a ubiquitously expressed protein that binds with SMC1A/B to form the legs of the isosceles triangle commonly used to represent the closed heterotrimeric cohesin ‘ring,’ with RAD21/REC8 forming the base.

In mouse models, homozygous *SMC3* loss^8,9^ or conditional *SMC3* depletion in oocytes^10^ results in embryonic lethality. In embryonic and adult mice, homozygous *SMC3* loss in the blood compartment causes myeloid-based hematopoietic failure.^11,12^ However, *SMC3* heterozygosity in mice is tolerated and associated with behavioral phenotypes, neuronal/synaptic differences in the cerebral cortex, decreased body weight, craniofacial dysmorphism, and changes in gene expression,^8,9^ suggesting the potential for a similar phenomenon in humans.^1^^3^

Missense variants and in-frame indels in *SMC3* have been identified among patients with mild/atypical Cornelia de Lange syndrome (CdLS).^13–15^ Typical (classic) CdLS is caused by constitutional or mosaic heterozygous loss-of-function (LoF) pathogenic variants in *NIPBL* and is distinguished from atypical CdLS, which is a spectrum, by greater overall severity with a more characteristic facial appearance and the presence of major malformations. Variable intellectual disability, behavioral abnormalities, hirsutism, growth failure, gastroesophageal dysfunction, and short first metacarpals are seen in both typical and atypical cases. *SMC3*-related CdLS is principally marked by intellectual disability, facial dysmorphism, microcephaly, and postnatal growth delay.^13–16^

The reported missense variants and in-frame indels in *SMC3* generally fall within important protein regions including the antiparallel coiled-coil, hinge, and head domains.^14^ Tested pathogenic variants appear to act as dominant-negative alleles in human cell lines, where they increase the affinity of SMC hinge dimers for DNA, promoting genome instability and impairing genomic spatial organization.^17^ *In silico* modeling is also consistent with a dominant-negative effect.^14^ Furthermore, proteomic analyses reveal that missense-mutated SMC3 proteins are incorporated into the cohesin complex normally but prompt dysregulation of the c-MYC transcription factor, a feature of CdLS in the early prenatal period.^18^ Finally, *SMC3* missense variants in CdLS patients confer a DNA methylation episignature grouping with that of other CdLS genes.^19^

There is considerable importance of allele type in other cohesin genes, for example *NIPBL* (missense in mild CdLS vs LoF in severe CdLS^20–22^) and *SMC1A* (missense in atypical CdLS vs LoF in *SMC1A*-related developmental and epileptic encephalopathy^23^). Thus, it has been proposed that the absence of *SMC3* predicted LoF (pLoF) variants in previously described CdLS cases could be due to those variants either causing a different phenotype or being lethal for the developing embryo.^24^ Somatic *SMC3* pLoF variants have been found in myeloid malignancies,^25^ including acute myeloid leukemia^26^ and Down syndrome acute megakaryoblastic leukemia.^27,28^ This is in line with LoF alleles in cohesin genes being a common event in several tumor types, not as initiating events but as subsequent drivers.^29^ Population genetic data suggest that germline *SMC3* pLoF variants are considerably depleted in the general population (https://gnomad.broadinstitute.org/gene/ENSG00000108055); however, the consequence of heterozygous germline pLoF variants in humans is unknown, as only a single case is documented in the literature.^15^

Here, we describe the clinical phenotypes of 13 individuals with germline pLoF variants in *SMC3*, including four frameshifts, four stop gains, four deletions, and one predicted damaging splice variant. Many, but not all, individuals have developmental delay, low growth parameters, and/or mild dysmorphism – reminiscent of atypical CdLS. Comparisons of quantitative growth and developmental data between pLoF carriers and published cases of *SMC3* pathogenic missense variants and in-frame indels reveal a genotype-phenotype correlation, with pLoF conferring milder – albeit overlapping – parameters. We also observe several individuals without growth or neurodevelopmental abnormalities and test multiple hypotheses to explain this incongruity. Our findings suggest that a broader class of undiscovered Mendelian conditions stemming from LoF variants in highly constrained genes likely remains uncatalogued owing to generic, variable, and/or incompletely penetrant phenotypes and will require large dataset analyses, deep phenotyping, and functional experimentation to solve.

## Subjects and Methods

### Case recruitment

Anonymized genetic and phenotypic information was collected, subjects were enrolled, and database searches performed under Boston Children’s Hospital IRB-approved protocol #00040134. Multiple sources were queried to identify individuals with predicted loss of function variants in *SMC3*: ClinVar (https://www.ncbi.nlm.nih.gov/clinvar/);^30^ GeneMatcher/Matchmaker Exchange (https://genematcher.org/);^31^ the Baylor Genetics, GeneDx, Indiana University School of Medicine Genetic Testing, and Invitae clinical laboratories; a database of patients at Boston Children’s Hospital;^32^ an in-house data portal built upon seqr^33^ and drawing from the Broad Institute Center for Mendelian Genomics and GREGoR Consortium projects; DECIPHER (https://www.deciphergenomics.org);^34^ and one previously-published individual with an *SMC3* nonsense variant referred for routine CdLS screening due to CdLS-like features and developmental delay.^15^

### Growth and developmental milestone comparisons

Growth measurements were converted to Z-scores using the British Growth Survey data (https://www.rcpch.ac.uk/resources/growth-charts). Statistical comparisons and plot generation were undertaken using R.

### Subject variant identification

Variants were identified via exome sequencing, microarray, or gene panel. These methods are listed for each case in Table S1, as is the confirmation status of variants by orthogonal methods.

### Gene annotations

RefSeq transcript NM_005445 (Gencode ENST00000361804.5), via (https://genome.ucsc.edu/) was used as the gene model for analyses. The Ensembl Variant Effect Predictor (https://useast.ensembl.org/Tools/VEP) was used to corroborate subjects’ variant nomenclature. Regional missense constraint for *SMC3* was based on gnomAD data and assessed at a threshold of p=0.001 as in https://www.biorxiv.org/content/10.1101/148353v1 and ^35^. The region of potential escape from nonsense mediated decay was considered as the final 55 nt of the penultimate exon of *SMC3* (codon 1176 and beyond). GRCh38/hg38 is the default genome build used unless specified otherwise.

### UK Biobank pLoF sequence variant curation

The 10-110575403-TGTGA-T splice site variant, present in 5 individuals in the UKBB, was removed from the list of *SMC3* pLoF variants in UKBB; this is on account of it not resulting in an alteration to the first 5 nt of the 5′ splice site.

### Copy-number variant detection among UK Biobank samples

UKBB *SMC3* deletions were called using GATK-gCNV with genomic interval selection, sample processing, and defragmentation performed as in (https://www.biorxiv.org/content/10.1101/2022.08.25.504851v1). In brief, exome sequencing reads were first mapped to a predefined set of genomic intervals curated to capture the exonic regions of canonical protein-coding genes, and coverage was collected across well-captured intervals for copy number (CN) inference. Samples with similar global read depth profiles were then clustered into batches to be jointly processed by GATK-gCNV, which outputs CNV calls across the set of well-captured intervals. These raw calls were subsequently defragmented, and to ensure that calls were not merged across high-quality CN=2 regions, CNVs were refragmented to preserve any CN=2 segments spanning at least 3 exome sequencing probes with quality score (QS) ≥100. Finally, the resulting callset was refined using the recommended QS, sample-level, and site frequency filters from (https://www.biorxiv.org/content/10.1101/2022.08.25.504851v1) along with a filter to remove CNVs covering fewer than 3 exons.

### Infertility cohort missense burden analysis

Comparisons were made to gnomAD v2.1.1 missense *SMC3* variants with CADD score >25 (Table S4) as assigned by the Variant Effect Predictor (https://useast.ensembl.org/info/docs/tools/vep/index.html). The cumulative allele frequency was transformed to a fraction with a denominator representing the mean number of alleles across these variants, for the purpose of chi-square analysis.

### Gene expression analyses

*SMC3* gene expression data were derived from Cancer Cell Line Encyclopedia (CCLE) samples, restricted to hematopoietic and lymphoid tumors, obtained via (http://xenabrowser.net). Nonsense, frameshift, or splice site variants were considered predicted loss-of-function (pLoF). Samples containing >1 *SMC3* variant were categorized by the most damaging variant class. Samples solely containing missense or silent *SMC3* variants were removed, as were samples lacking *SMC3* genotype or expression data.

Perturb-seq data were derived from Replogle et al.^36^ and obtained via (https://plus.figshare.com/articles/dataset/_Mapping_information-rich_genotype-phenotype_landscapes_with_genome-scale_Perturb-seq_Replogle_et_al_2022_-_commonly_requested_supplemental_files/21632564/1) and (https://gwps.wi.mit.edu/). Briefly, this Perturb-seq experiment involved single-cell sequencing following pooled, multiplexed CRISPR interference (CRISPRi) in three parallel approaches, each using a different CRISPRi guide library: K562 (chronic myelogenous leukemia, female) cells receiving a genome-wide guide library against all expressed genes and harvested on day 8 after transduction; K562 cells receiving a DepMap essential gene guide library and harvested on day 6 after transduction; and hTERT RPE1 (retinal pigment epithelium, female) cells receiving a DepMap essential gene guide library and harvested on day 7 after transduction. Each library vector construct encoded two guides (sgRNAs) per target gene.

*Smc3*^+/-^ mouse cortex differentially expressed genes from RNA sequencing were obtained from Fujita et al.^8^ *Nipbl*^+/-^ whole brain RNA sequencing data were obtained from Kean et al.^37^ In comparisons of these datasets, a denominator of 30,686 genes (see Fig. 3d) was established from among the 55,228 total genes mapped in Kean et al.^37^ (GEO entry: https://www.ncbi.nlm.nih.gov/geo/query/acc.cgi?acc=GSE203014) based on those having 10 or more reads across all 22 samples, which was the cutoff used in the original analysis.

Statistical analyses were conducted in R. Key gene lists are in Table S5.

### DNA methylation analysis

DNA from blood was subjected to array-based methylation analysis via the Manchester EpiPro project (https://mft.nhs.uk/nwglh/test-information/rare-disease/epipro-project/). The genome-wide DNA methylation profile was compared to the EpiSign Knowledge Database, a collection of DNA methylation signatures specific for rare disorders, as in Aref-Eshghi et al.^19^

## Results

### Case series and subject phenotypes

Thirteen individuals with *SMC3* predicted loss of function (pLoF) variants were identified. Most subjects underwent genetic testing as part of routine clinical assessment for growth retardation, developmental delay, and/or dysmorphic features. In only three cases was there clinical suspicion of CdLS. Some cases were identified via diagnostic laboratories, research repositories, hospital-based databases, and GeneMatcher. Subjects possessed frameshift indel, deletion, stop gain, or splice variants (Tables 1, S1; Fig. 1). Of these 13 variants, five were *de novo*, one was maternally inherited, two were paternally inherited, and six were of unknown inheritance. Adjudication of the nine SNVs using an advanced LoF curation framework (https://www.medrxiv.org/content/10.1101/2023.03.08.23286955v1) flagged two as potentially not having a LoF effect (one splice variant and one escape from nonsense mediated decay) (Table S1). Missense variants were specifically excluded, as missense variant effects (e.g., hypomorphic vs nullimorphic vs dominant negative) are unable to be reliably predicted.

**Figure 1.**
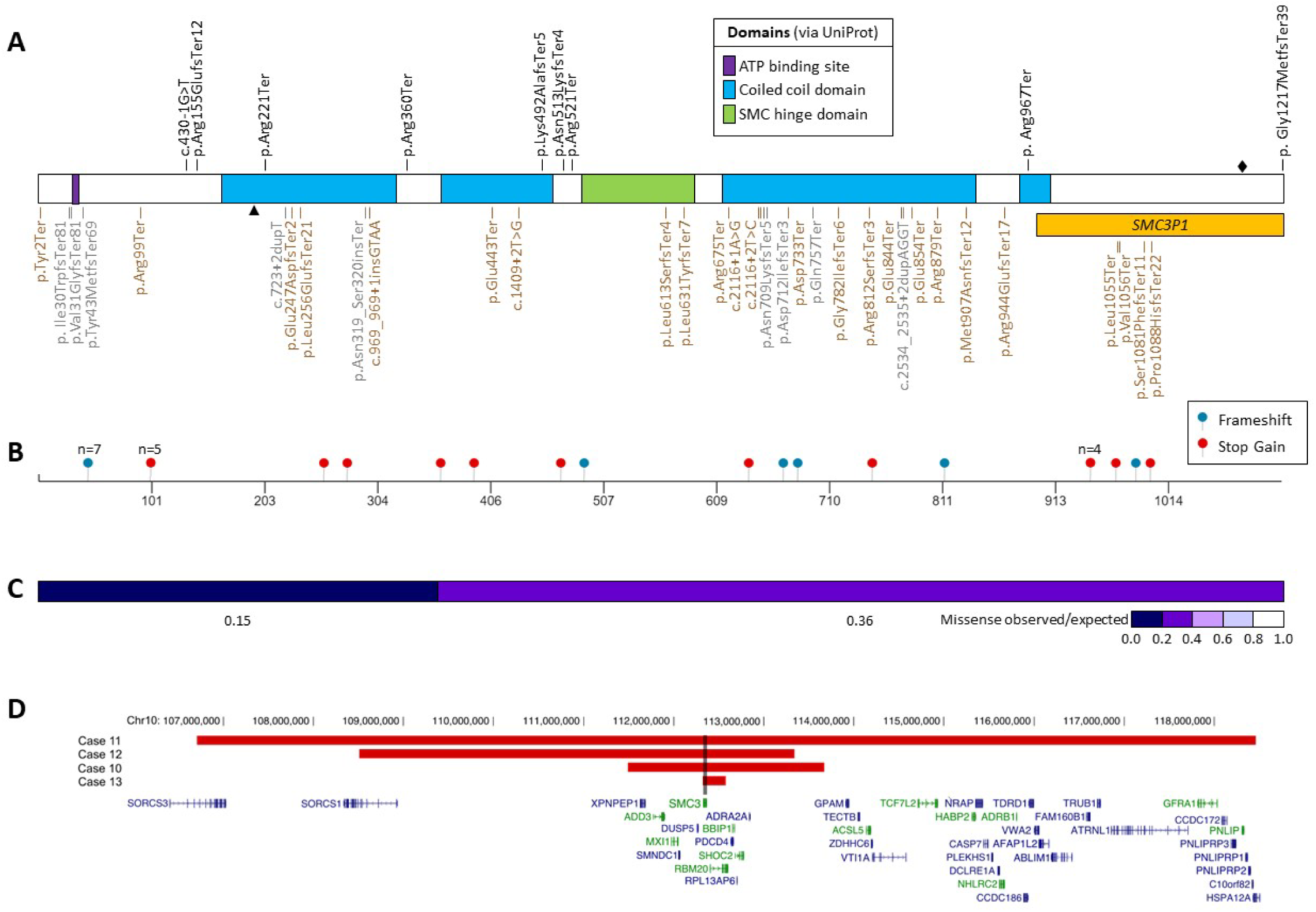
SMC3 predicted loss-of-function (pLoF) variants. **a.** pLoF simple nucleotide variants (SNVs) mapped to the SMC3 protein sequence. Subject cases are in black, top. The c.430-1G>T splice variant immediately precedes exon 8, which if skipped would result in a shift of reading frame, however splicing may be rescued (see text). The p.(Gly1217MetfsTer39) variant is in the final exon and is predicted to escape nonsense mediated decay. gnomAD pLoF variants passing quality filters are in grey, bottom. UKBB pLoF variants are in brown, bottom. Domains via UniProt.^73^ Arrowhead denotes the C-terminal end (equivalent to codon 211) of the minor transcript described in Fig. 3. Diamond denotes the point after which nonsense mediated decay may be escaped (codon 1176). The 3′ region homologous to the *SMC3P1* pseudogene (codons 974-1217) is in orange. **b.** Somatic *SMC3* pLoF SNVs among 13,714 tumors from the NCI GDC collection (https://portal.gdc.cancer.gov/genes/ENSG00000108055). **c.** Regional missense constraint over *SMC3*, via an analysis of gnomAD. The transition point is in codon 390 (exon 13 of 29). **d.** Case deletions (based on genome build GRCh37/hg19). Of the OMIM phenotype-associated genes in this region (green), none are dominant haploinsufficient disease genes for disorders that include developmental delay or dysmorphic features.

Information on individual patients’ clinical features can be acquired by contacting the corresponding authors. This information will be included in a future peer-reviewed manuscript but is prohibited by medRxiv. No feature was shared by all subjects, however the features present in more than half of subjects included developmental delay and/or intellectual disability, low growth, and facial dysmorphism. The majority of the typical CdLS facial features and associated malformations were rare or absent in our patient cohort. No low anterior hairline or major limb malformations were reported. Only two individuals had a long/featureless philtrum, one had ptosis, three had confirmed dental anomalies, and one had downturned corners of the mouth. Three individuals had synophrys, three had arched eyebrows, and five had long eyelashes. Three individuals had brachycephaly, three had a thin upper lip, and three had a depressed nasal bridge. Four individuals had cardiac malformations, and two experienced gastroesophageal reflux (GER). Hirsutism was described in three cases. Interestingly, five out of thirteen individuals had a degree of micrognathia/retrognathia. Intellectual disability was present in four individuals and learning disability in an additional one. A delay in reaching developmental milestones was detected in nine individuals. Autistic features were present in three individuals. Photographs were not available for any subject with dysmorphic features.

We compared standardized postnatal growth parameters and age at reaching developmental milestones between carriers of *SMC3* pLoF variants and 15 cases with pathogenic missense variants or in-frame indels in *SMC3* as previously published.^14^ A difference in standardized birth weight, birth head circumference, postnatal (i.e. at time of enrollment) weight, and postnatal head circumference was detected between the two groups, pLoF variants being associated with less pronounced growth delay than missense/in-frame indel variants, however these differences did not reach a significance threshold of p≤0.05 (Fig. 2a). When comparing the age at achieving developmental milestones, the patients with pLoF variants were able to sit unaided, walk unaided, and speak at a younger age than patients with missense/in-frame indels, although only the difference in age at sitting was significant (p=0.023) (Fig. 2b). pLoF subjects’ growth and developmental milestones were each delayed compared with the population mean in growth and developmental data (Fig. 2). Thus, in general *SMC3* pLoF variants are associated with a milder phenotype than are missense variants; however, the spectra are variable and appear to overlap.

**Figure 2.**
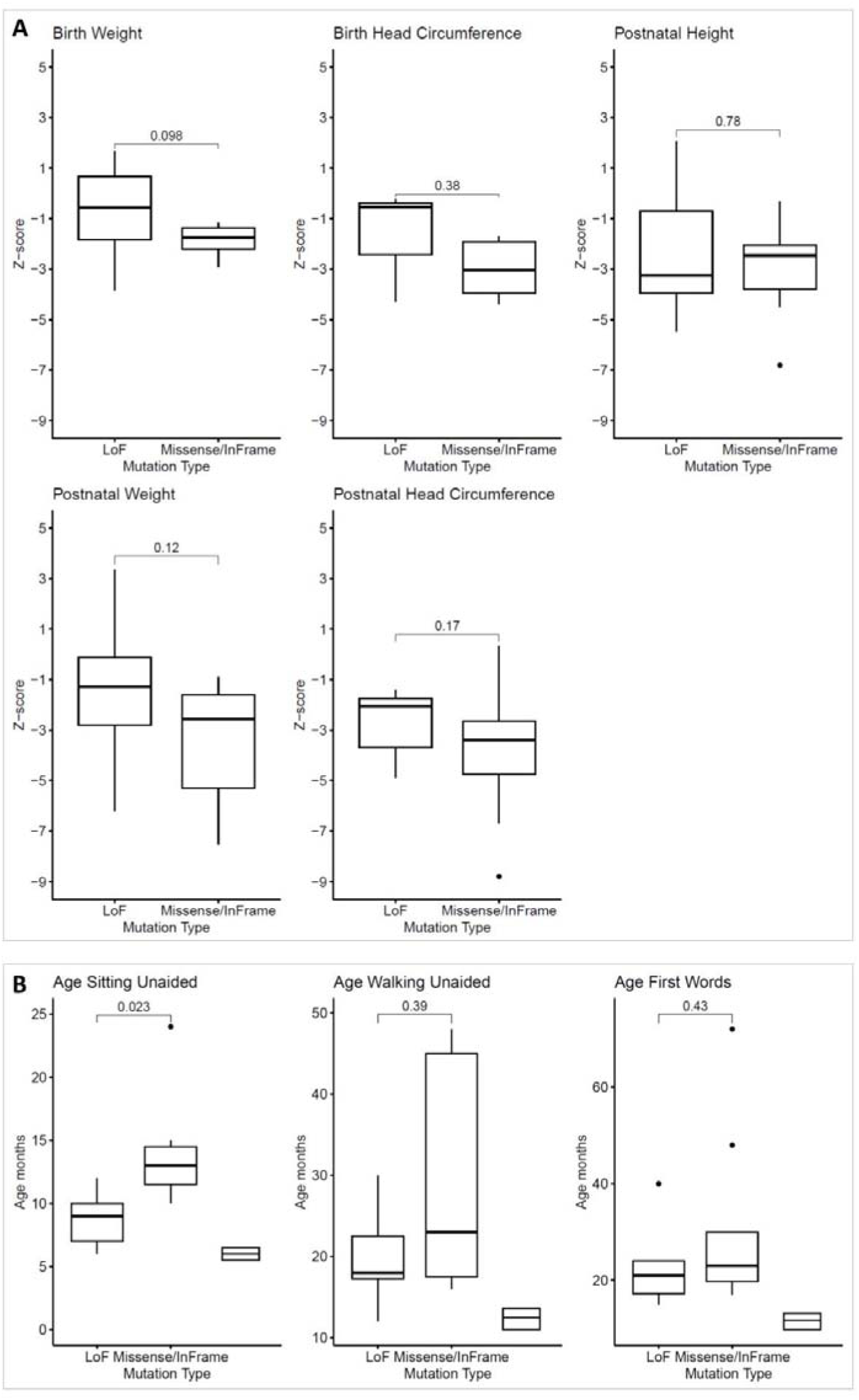
Growth and developmental milestones associated with *SMC3* pLoF compared with pathogenic missense/in-frame indels. **a.** Growth measurements adjusted for age and sex (Z-score) are shown for *SMC3* predicted loss-of-function (LoF) and missense/in-frame indel cases. ‘Postnatal’ measurements are at the time of study enrollment/sampling. **b.** Age at which developmental milestones were reached for the two categories of *SMC3* variants (LoF vs pathogenic missense/in-frame indel). 25^th^/50^th^/75^th^ centiles of population data (Denver II, via DECIPHER) are shown at the right of each figure in (**b**). Missense/in-frame indel data were obtained from ^14^. Plots are standard boxplots. Statistical comparisons are the Mann-Whitney U test (Wilcoxon).

**Figure 3.**
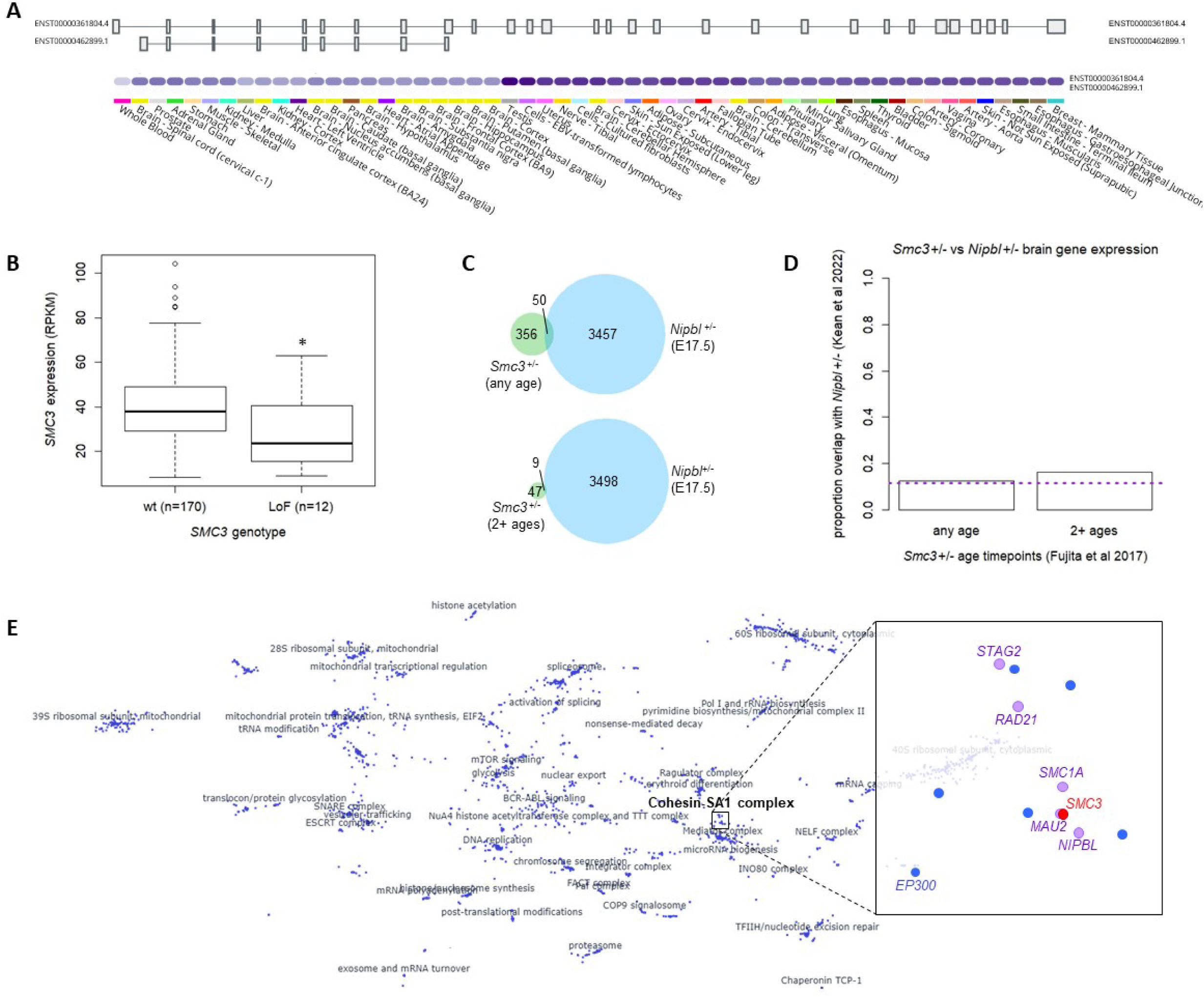
Gene expression in wild-type and *SMC3*^+/-^ tissues. a. GTEx data demonstrate that only a single, full-length *SMC3* isoform (ENST00000361804.4) is expressed at an appreciable level in any adult human tissue represented (https://gtexportal.org/home/gene/SMC3). **b.** pLoF *SMC3* variants (LoF; nonsense, frameshift indel, splice site) in hematopoietic and lymphoid tumors correlate with decreased *SMC3* gene expression, to a mean of 71.9% of wild-type (wt) (p=0.019, Mann-Whitney U test). Data derived from the Cancer Cell Line Encyclopedia (CCLE) (http://xenabrowser.net). **c-d.** Comparison of genes with altered expression in early postnatal *Smc3*^+/-^ mouse cortex^8^ and those altered in a late prenatal *Nipbl*^+/-^ mouse whole brain.^37^ **c.** 50 of 406 (12.3%) differentially expressed genes (DEGs) any age (P1, P3, P7, P14, P21) in *Smc3*^+/-^ mice overlap with 3,507 DEGs in E17.5 *Nipbl*^+/-^ mice (top). 9 of 58 (15.5%) DEGs in 2+ ages in *Smc3*^+/-^ mice overlap with those same 3,507 genes (bottom). **d.** Neither of these is significantly different from chance (3,507/30,686 total genes; 11.4%) (any age p=0.632, chi-squared test of proportions; 2+ ages p=0.441). See Methods for derivation of denominator of 30,686 genes and Discussion for a commentary on methodological differences between the two models. **e.** Minimal displacement embedding of pseudobulk-averaged, z-normalized expression profiles from a genome-wide Perturb-seq experiment in K562 CML cells^36^ (plot generated via https://gwps.wi.mit.edu/). Each dot is one of 1,973 genes with strong transcriptomic signatures. The *SMC3* knockdown transcriptional disturbances group tightly with those of other cohesin genes (pink) and even an epigenetic regulator occasionally mutated in CdLS-like patients (*EP300*).

Interestingly, several subjects had other phenotypes with potential biological links to *SMC3* (see Discussion) including bone marrow failure or leukopenia, acute myeloid leukemia (AML), and Coats retinal telangiectasis, however these were limited to a single case each. Additional phenotypic and laboratory details, where available, are in the Supplemental Note.

### SMC3 mutational constraint

We were intrigued by subjects who were nonpenetrant for, or had mild presentations of, the above growth and developmental phenotypes given initial metrics suggesting mutational intolerance in this gene. Thus, we sought to further characterize LoF constraint in *SMC3*. Consistent with *in vitro*^38,39^ and model organism studies^9,10,40,41^ indicating *SMC3* is an essential developmental gene, biallelic pLoF variants have never been described in a human being, including our cases.

Regarding heterozygous variants, the gnomAD v2.1.1 (https://gnomad.broadinstitute.org/) probability of LoF intolerance (pLI) score (evidence for depletion of functional variation from expectation within each gene) and the loss-of-function observed/expected upper bound fraction (LOEUF; a more continuous metric that directly measures the ratios of observed to expected variation) both place *SMC3* among the group of genes predicted to be most highly intolerant to LoF variants in a population cohort of adults without severe pediatric disease (pLI = 1; expected/observed ratio of SNV pLoF variants passing filters = 79.5/0; LOEUF 0.04).^42^ In agreement with this, an estimation of the probability of haploinsufficiency (pHaplo) of *SMC3* based on a machine learning model trained on a broad case-control CNV burden analysis in >950,000 individuals and gene-level features was 0.998 (max score = 1.00);^43^ of interest, the probability of triplosensitivity (pTriplo) is also very high (0.999). A review of multiple additional databases of control and affected subjects revealed rarity and considerable apparent selection against such alleles (Table S2): No pLoF structural variants are present in gnomAD-SV, and our own CNV analysis of UK Biobank (UKBB) exome data identified only two deletions involving *SMC3* among 196,869 subjects (Fig. S2). pLoF sequence and structural variants are similarly rare or absent in the Database of Genomic Variants (DGV) and DECIPHER (Table S2).

To gain insights into the few heterozygous *SMC3* pLoF SNV carriers in control populations, who would presumably be nonpenetrant or mildly affected, we cataloged these alleles in gnomAD and the UK Biobank (UKBB) (Fig. 1a; Table S3). The overall frequency of pLoF variants in gnomAD is 6.37E-5 (1/15,699) and in UKBB is 3.32E-5 (1/30,102), and all are singletons. There was neither a strong indication of mosaicism (e.g., low allele balance) nor an age-dependent increase in allele frequency suggestive of clonal hematopoiesis among gnomAD pLoF variants (Fig. S1). Adjudication of the nine gnomAD pLoF variants using the LoF curation framework described above flagged two as not having a LoF effect and an additional two as uncertain or potential technical artifacts (e.g., homopolymer repeat region, lack of read data) (Table S3).

Mapping case pLoF SNVs to the *SMC3* coding sequence suggested possible clustering toward the 5′ half of the transcript (Fig. 1a). To identify gene-level features that would support the veracity and/or biological meaning of such a cluster, we first referenced GTEx (https://gtexportal.org/home/gene/SMC3) and found that the full-length, canonical transcript is the only isoform expressed at an appreciable level in any adult human tissue (Fig. 3a). Further supporting this being the sole isoform, normalized by-exon expression as measured by pext^44^ (obtained via gnomAD) was homogeneous (score=1) across all exons in every GTEx tissue. Next, we investigated whether the existence of *SMC3P1*, a homologous pseudogene at 2q11.2 that is likely the result of retrotransposition of approximately the last 5 exons of *SMC3* (Fig. 1a), might be impairing variant mapping in that region resulting in an artifactual depletion of 3′ variants. However, per-base mean depth of coverage among gnomAD exomes and genomes is similar across the entirety of *SMC3* (https://gnomad.broadinstitute.org/gene/ENSG00000108055?dataset=gnomad_r2_1), and *SMC3* pLoF variants in gnomAD/UKBB (Fig. 1a) and somatic *SMC3* pLoF variants in tumors from the NCI GDC collection (Fig. 1b) show an even spread across the gene. Finally, we calculated regional missense constraint scores across *SMC3* (Fig. 1c); while constraint scores are slightly higher at the 5′ end, the entire gene is substantially missense constrained (o/e = 0.28 (0.25 - 0.32); Z=6.4). These data argue against any meaningful clustering of pLoF SNVs.

### Potential explanations for mild phenotypes and nonpenetrance

Having made a strong case for intolerance to heterozygous LoF of *SMC3*, we continued to seek to explain cases with mild or nonpenetrant developmental and/or growth phenotypes. One potential explanation is mosaicism. Thus, we assessed inheritance and allele balance among our enrolled cases. Variants were inherited in three cases and thus by definition germline mutations in those probands. No case variants were signed out by clinical labs as having allele fractions suggestive of mosaicism. Furthermore, in one subject lacking growth or developmental delay, we identified the mutant allele in three separate tissues (bone marrow, skin biopsy, and blood) with a near-50% allele balance (Supplemental Note).

Another potential explanation for mildly- or un-affected individuals with variants in considerably haploinsufficient genes is that they are dominant infertility/subfertility genes. Indeed, testis and ovary are the adult human tissues with highest relative expression of *SMC3* (https://gtexportal.org/home/gene/SMC3). Yet, *SMC3* has never been identified as a human infertility gene, including the most recent large study of primary ovarian insufficiency,^45^ and we identified one maternally transmitted *SMC3* pLoF variant. We tested an alternative hypothesis that heterozygous LoF of *SMC3* might cause male infertility. We evaluated WES data from >1,000 individuals with non-obstructive azoospermia (NOA) in the GEMINI Phase I cohort^46^ and >1200 additional exomes in a Phase II cohort (some of which were normozoospermic male controls and female infertility cases). No *SMC3* pLoF variants and only one missense variant with CADD score ≥25 were identified (Table S4). Additionally, we assessed rare (gnomAD MAF <1%) coding variants in the Male Reproductive Genomics (MERGE) cohort,^47^ consisting of 2,100 exomes of infertile men with severe oligo-, crypto-or azoospermia. One pLoF and three missense variants with CADD ≥25 were identified (Table S4). MERGE missense variants were not enriched when compared with gnomAD variants with CADD ≥25 (p=0.612, 2-sample chi-squared test with continuity correction). Finally, among our cases were two paternally inherited variants.

### Functional effect

We next sought to determine whether pLoF *SMC3* variants act as true LoF alleles and have broader functional genomic effects. As subject cell lines and RNA were not available, pLoF *SMC3* variants were identified among hematopoietic and lymphoid tumors from the Cancer Cell Line Encyclopedia (CCLE) with available RNA sequencing data; these led to a significant decrease in *SMC3* gene expression compared to tumors wild-type for *SMC3* (p=0.019, Mann-Whitney U test) (Fig. 3b).

To determine whether decreasing *SMC3* expression is likely to effect a molecular (transcriptomic) signature and to what extent this matches that of other CdLS/cohesinopathy genes, we analyzed Perturb-seq data from Replogle et al^36^. This resource employed multiplexed CRISPR interference followed by single cell RNA-sequencing. In a genome-wide experiment in K562 chronic myelogenous leukemia cells, cells receiving *SMC3*-targeting guide RNAs reduced *SMC3* expression to 68.5% residual and exhibited a transcriptomic signature of several hundred up- and down-regulated genes (Table S6). The expression profile of *SMC3* knockdown was highly correlated with the knockdown profiles of other cohesin ring components (*SMC1A*, *RAD21*, *STAG2*) and the cohesin loading machinery (*NIPBL*, *MAU2*) (Fig. 3e). This correlation held true in separate Perturb-seq experiments targeting only essential genes in K562 or retinal pigment epithelium (RPE1) cells (Fig. S3). The data were also able to show a threshold above which *SMC3* knockdown – despite cohesin being involved in chromosome segregation – does not result in chromosome instability, similar to other cohesin genes (Table S6) and previous studies involving *SMC3*.^48^

*Smc3*^+/-^ mice were previously shown to possess differences in P1-21 cortical gene expression,^8^ in addition to behavioral differences. Despite substantial methodologic differences – for example age at tissue harvest, which is associated with significant changes in cohesin level^8^ – we ventured to compare these differentially expressed genes to those observed in a study of E17.5 *Nipbl*^+/-^ mouse whole brain, a model of typical CdLS.^37^ No significant overlap was observed (Fig. 3c).

Finally, we sought to determine whether *SMC3* pLoF variants result in changes to the epigenome matching those of previously-described subjects with CdLS including individuals with missense and in-frame indel variants in *SMC3*, although largely driven by other CdLS genes.^19^ Methylation analysis of blood DNA from one available subject (p.Arg360Ter) did not match that episignature (Fig. 4).

**Figure 4.**
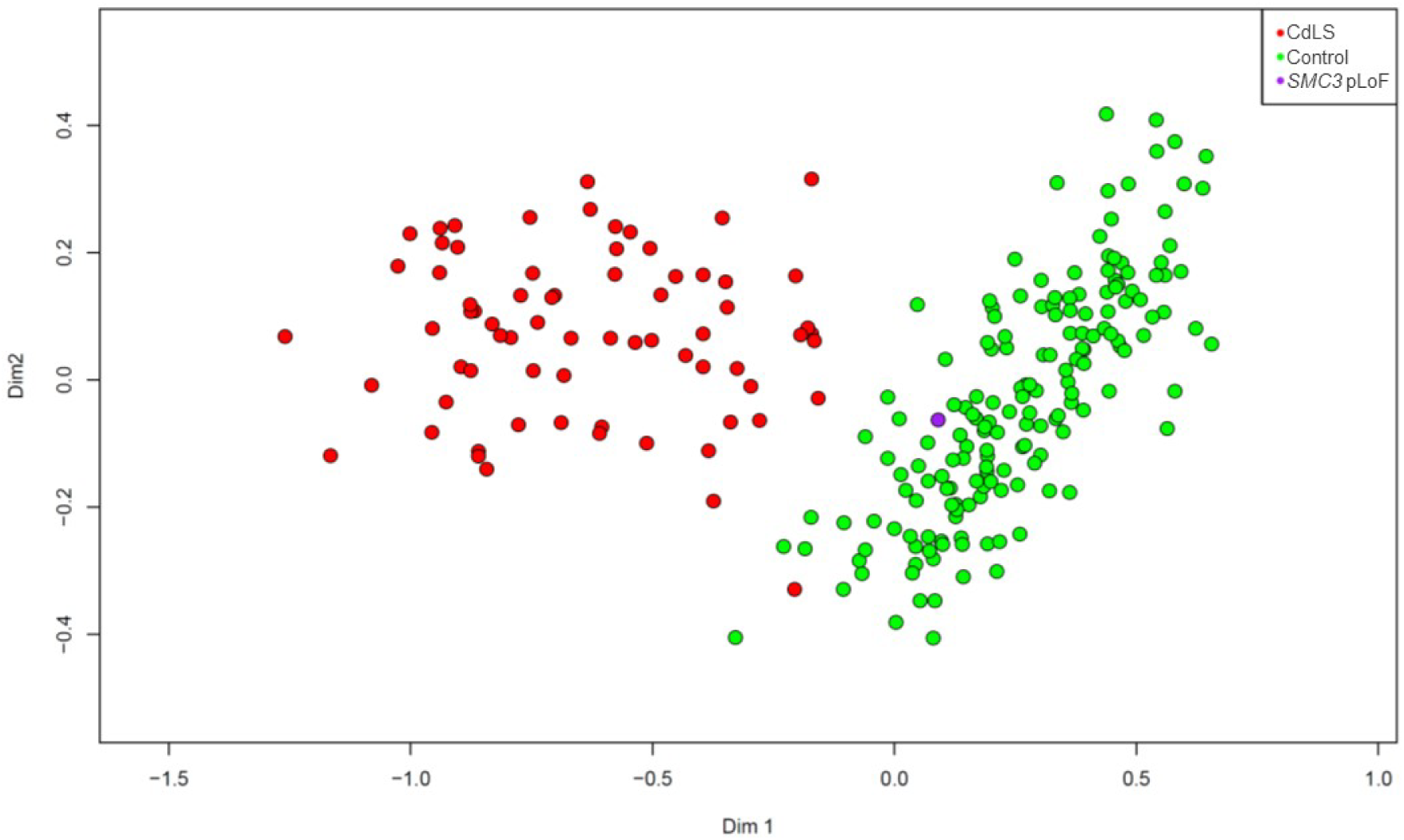
Global DNA methylation pattern of *SMC3* pLoF compared with that of CdLS. Multidimensional scaling (MDS) plot of global methylation signature (episignature) of blood-derived DNA. A *SMC3* pLoF subject (p.Arg360Ter, purple) does not plot with individuals clinically diagnosed with CdLS (red), instead plotting among the control population (green).

**Figure 5.**
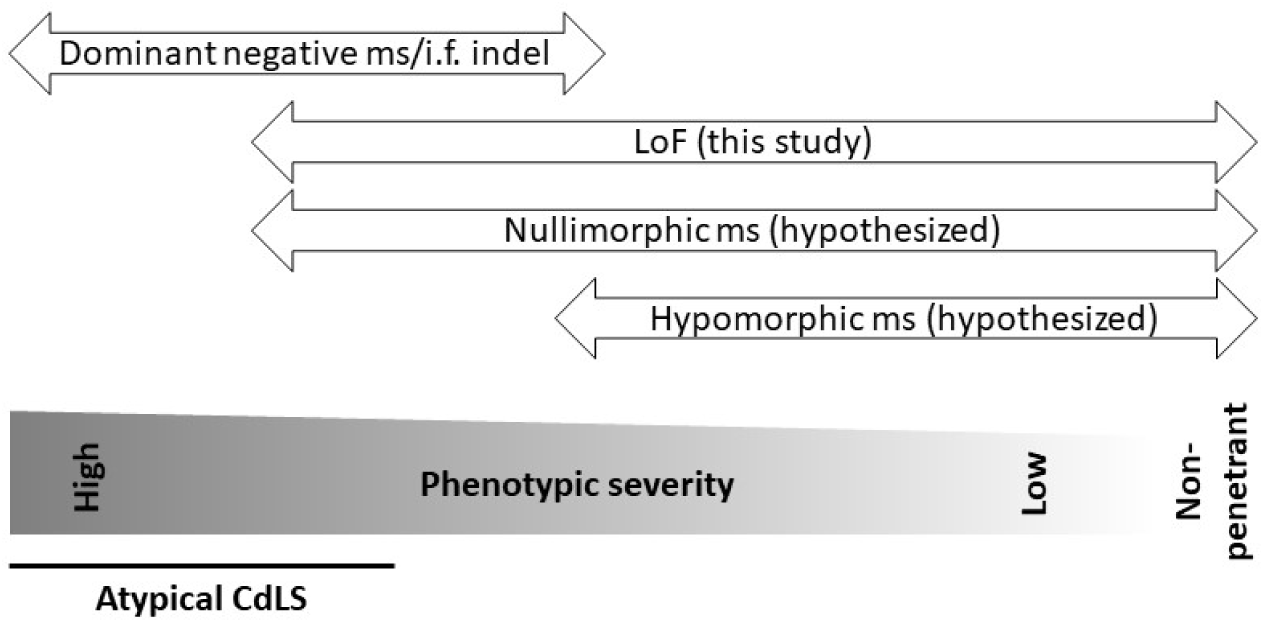
Hypothesized phenotypic spectra of *SMC3* variant types. Missense (ms) /in-frame (i.f.) indel variants, via an apparent dominant negative mechanism as supported by prior literature, carry the most severe phenotype, with many patients appearing to have atypical Cornelia de Lange syndrome (CdLS). Loss-of-function (LoF) variants carry a milder phenotype, with only some individuals being recognized to have CdLS and some individuals lacking key phenotypic features (growth retardation, developmental delay, characteristic facial dysmorphism). We hypothesize that nullimorphic ms and hypomorphic ms variants also exist.

## Discussion

While heterozygous *SMC3* missense and in-frame indel variants are a cause of atypical Cornelia de Lange syndrome (CdLS), and somatic *SMC3* predicted loss-of-function (pLoF) variants are found in cancers, the consequence of germline *SMC3* LoF variants in humans has remained solely in the realm of speculation. The present study was aimed at resolving this question.

### Clinical phenotype

The *SMC3* pLoF cases presented here demonstrate that this variant type also contributes to a disease phenotype. Published *SMC3* missense and in-frame indel cases with atypical CdLS demonstrate growth retardation, developmental delay, and characteristic facial features.^14^ We found that pLoF patients share these features, although by quantitative comparisons appear on average to have a milder – although overlapping – phenotype than those with missense/in-frame indel variants and would for the most part not be considered to have CdLS, which might explain the near-absence of *SMC3* pLoF variants among previously sequenced CdLS cohorts (Fig. 5). Furthermore, the phenotype is variable, with some subjects showing more severe growth or developmental delays and others being nonpenetrant for these key features. Of note, *Smc3*^+/-^ mice have demonstrated neuronal, behavioral, growth, and craniofacial features.^8,9^

The pathogenesis of CdLS in previously-reported *SMC3* missense/in-frame cases has been suggested to be dominant negative, involving aberrant pairing of mutant SMC3 with other components of the cohesin ring.^17,18^ Our data suggest that, by contrast, a relative shortage of SMC3 has a less severe impact on cohesin function, which suggests that allele-specific therapy to knock down dominant negative alleles – even completely – should be further explored. Nullimorphic and hypomorphic *SMC3* missense variants likely also exist and will be identified as such with further study (Fig. 5).

### Deletions involving SMC3

In the four patients with deletions at 10q25.2, *SMC3* is not the only gene involved in the rearrangement (Fig. 1d) and it is therefore possible that haploinsufficiency for other genes in the region is causative for some aspects of their phenotype. A small number of genes in these intervals have been associated with dominant human disease, among which only heterozygous *SHOC2* and *SMC3* pathogenic variants cause dysmorphic features and developmental delay; however, the only known pathogenic *SHOC2* variant which causes Noonan-like syndrome with loose anagen hair (OMIM #607721) is missense. Previously reported 10q25.2 deletions detected by cytogenetic methods do not seem to be associated with CdLS-like facial features.^49,50^

Missense variants in *RBM20*, which is deleted in all four deletion cases, are associated with a form of dilated cardiomyopathy.^51^ No patients in our study with deletions encompassing *RBM20* had any sign of cardiomyopathy. The small deletion which only contains *SMC3* and *RBM20* in one subject was of particular interest, as it was also detected in the subject’s father who was reported to be healthy with no mention of heart disease.

### Potential explanations for mild/nonpenetrant cases despite LoF constraint

Analyses of multiple population genomic analyses suggested that *SMC3* is substantially LoF-, missense-, and even duplication-constrained. Given substantial LoF constraint, it was then potentially surprising that we identified some individuals as having milder features as well as pLoF individuals in gnomAD and UKBB (albeit at a low allele frequency). Of note, constraint metrics are indicative of overall selective pressure rather than observable phenotypic severity,^52^ and the above scenario has been described for other, similarly LoF-constrained disease genes (e.g. *ASXL3, ARID1B* and *AUTS2*).^53^ Nonetheless, we sought to investigate potential alternative explanations: Obligatory postzygotic mosaicism is seen in disorders for which germline pathogenic variation is lethal.^54^ Mosaicism restricted to the blood lineage may be related to clonal hematopoiesis in aging,^55^ which can be caused by somatic variants in a number of cohesin and related genes (e.g. *RAD21*, *SMC1A*, *STAG2*, *CTCF*)^56^. However, somatic *SMC3* pathogenic variants have not previously been associated in isolation with clonal hematopoiesis,^12^ and we found no evidence for mosaicism in our cases or in the few pLoF variants in gnomAD.

In mice, sufficient SMC3 protein is required in oocytes for early embryonic development,^10^ and heterozygous depletion of *SMC3* in female mice has deleterious impacts on both the integrity and transmission of zygotic chromosomes.^9,10,12^ Furthermore, conditional knockout of *HDAC8*, an SMC3 recycling factor, causes subfertility.^57^ Finally, several cohesin or cohesin-related genes have been implicated in infertility in humans (*REC8*, *SMC1B*, *STAG3*, *SGO2*).^58^ Yet, there is no published evidence of *SMC3* being involved in female infertility and our own analyses identified only rare *SMC3* pLoF variants among male infertility (azoospermia) cases and no statistical enrichment of predicted damaging missense variants.

### Case-control phenotype associations

*SMC3* deletions or pLoF (frameshift, stop gain, splice site) variants have not previously been found to be associated with disease in large case-control studies. For example, there were no control deletions or case deletions <1Mb in a large study generating a copy-number variant ‘morbidity map’ of individuals with developmental delay,^59^ and only one protein truncating variant (included in the present manuscript) was found among DDD study data.^60^ The UK Biobank (UKBB) demonstrates only one moderately significant phenotypic association for pLoF variants (mean corpuscular volume) (https://app.genebass.org/gene/ENSG00000108055)^61^. Intriguingly, in an analysis of >31k individuals with neurodevelopmental phenotypes and their family members, *de novo* variants in *SMC3* were found to be associated with developmental delay (false discovery rate = 3.46E-7), although driven mostly by missense variants.^62^ The lack of phenotypic associations with *SMC3* pLoF variants in large cohorts could be on account of a lack of true association and/or lack of power owing to the rarity of these variants.

### The possibility of pleiotropic phenotypes

Several patients had intriguing ‘other’ phenotypes with rational potential links to *SMC3*: 1) One subject had cytopenias and somewhat low telomere length, while being otherwise remarkably healthy with no intellectual disability, facial dysmorphisms, or other phenotypes consistent with CdLS (Table 1, Supplemental Note). Another subject also had leukopenia. This is of interest on account of poor hematopoietic replicative potential across serial transplantation experiments in *Smc3*^+/-^ mice and experiments showing that *SMC3* mutant human cells are out-competed by normal cells;^12^ however, others found increased self-renewal.^11,63^ Poor hematopoietic renewal could also result from telomere dysfunction, and some basic science studies potentially implicate cohesin in telomere biology,^64^ although CdLS patients (mostly with *NIPBL* variants) do not have short telomeres.^65^ Of note, the subject with low telomere length also had a VUS in *RTEL1*, a telomere biology disorder gene (OMIM 608833), that is absent from gnomAD and affects a conserved amino acid, although it was transmitted from his father with normal telomere length. 2) A subject with acute myeloid leukemia is also interesting on account of *SMC3* variants seen as secondary hits in this and other types of malignancy.^29^ 3) Finally, another subject had bilateral Coats disease, an ultra-rare, idiopathic retinal telangiectasia leading to intra-retinal and subretinal exudates.^66,67^ Syndromic associations with Coats disease include facioscapulohumeral muscular dystrophy (FSHD); approximately one percent of FSHD1 patients have Coats disease, ∼1,000 times higher than the general population.^68^ FSHD1 is caused by an autosomal dominantly inherited loss of D4Z4 repeats on chromosome 4q35, contracting the array from 11-100 to 1-10 repeats and promoting *DUX4* retrogene mis-expression.^68,69^ The remaining five percent of FSHD cases (FSHD2) may be attributed to genetic pathogenic variants that lead to repeat-independent hypomethylation of D4Z4 in genes such as *SMCHD1* (involved in genome organization) and *DNMT3B* (a chromatin modifier), also prompting mis-expression of *DUX4*.^70,71^ Based on the involvement of *SMCHD1* and *DNMT3B* in FSHD, perhaps other chromatin modifiers or cohesin genes such as *SMC3* may impact 4q35 chromatin decompaction, causing *DUX4* mis-expression and leading to FSHD. Unfortunately, additional functional testing to explore this hypothesis (D4Z4 repeat length, D4Z4 methylation, 4qA vs B haplotype, telomere length, and genome-wide methylation analyses) were unable to be performed for this subject.

**Table 1.** Details of heterozygous *SMC3* predicted LoF variants. VUS, variant of uncertain clinical significance. AOH, absence of heterozygosity. Pat, paternal. Mat, maternal. u, unknown/unreported inheritance. Variant nomenclature is based on GenBank accession NM_005445.4 (MANE Select).

| Case no. | 1 | 2 | 3 | 4 | 5 | 6 | 7 | 8 | 9 | 10 | 11 | 12 | 13 |
| --- | --- | --- | --- | --- | --- | --- | --- | --- | --- | --- | --- | --- | --- |
| Variant type | splice | frameshift | nonsense | nonsense | frameshift | frameshift | nonsense | nonsense | frameshift | whole gene del | whole gene del | whole gene del | whole gene del |
| Variant | c.430-1G>T p.(?) | c.461dup<br>p.(Arg155GlufsTer12) | c.661C>T<br>p.(Arg221Ter) | c.1078C>T<br>p.(Arg360Ter) | c.1474_1478del<br>p.(Lys492AlafsTer5) | c.1539del<br>p.(Asn513LysfsTer4) | c.1561C>T<br>p.(Arg521Ter) | c.2899C>T<br>p.(Arg967Ter) | c.3646_3647dup<br>p.(Gly1217MetfsTer39) | arr[GRCh37]<br>10q25.1q25.2(111490480_113668388)x1 | arr[GRCh37]<br>10q25.1q25.3(106709945_118460100)x1 | arr[GRCh37]<br>10q25.1q25.2(108508297_113338552)x1 | arr[GRCh37]<br>10q25.2(112323003_112579895)x1 |
| Inheritance | *de novo* | u | mat | pat | *de novo* | u | u | *de novo* | u | *de novo* | *de novo* | not mat | pat |
| Other |  |  | VUS dup<br>1q22q23.1, pat<br>*KMT2D* variant |  |  | Several other<br>variants, AOH |  | Short telomeres,<br>*RTEL1* VUS |  | VUS dup at 12q |  | Inherited chr16 del | Inherited inv(10) |

### Functional effect of heterozygous SMC3 pLoF variants

We analyzed hematologic and lymphoid transcriptome data, which suggested that pLoF *SMC3* alleles act as LoF alleles at the RNA level. This is in line with *in vitro* data showing the p.(Arg245Ter) variant causes loss of cohesin assembly.^63^ Analysis of Perturb-Seq data demonstrated impeccable grouping of the transcriptomic signature from *SMC3* knockdown with those of other cohesin ring and cohesin loading genes. An analysis of mouse brain transcriptome data failed to find significant overlap between differentially expressed genes in *Smc3*^+/-^ and *Nipbl*^+/-^ mice (a model of classic CdLS); however, substantial methodological differences existed between the construction, tissue sampling, RNA preparation, and analyses of these two models. DNA methylation in human blood of an *SMC3* pLoF case did not cluster with CdLS subjects on a robust epigenetic analysis platform, yet future work is needed to identify whether *SMC3^+/-^*LoF yields its own signature, either distinct from or an attenuated version of the CdLS signature.

### Summary

We demonstrate that *SMC3* pLoF variants are depleted at the population level, yet survivable, and provide evidence that they are associated with developmental phenotypes. In most cases it is associated with mild-moderate developmental delay, growth deficiency, and/or facial dysmorphism, although not all individuals displayed these features. On average these variants bear a phenotype milder than but overlapping with that of *SMC3* missense/in-frame indel variants present in CdLS cohorts.

While incomplete penetrance for a given disease feature – which we observed in our cohort, is still consistent with substantial population-scale LoF constraint,^52^ we nonetheless tested alternative explanations for this including mosaicism and infertility phenotype, neither of which was confirmed. Thus, an overall moderate-severity, but variable, phenotype is the apparent effect of these variants. Variants in *SMC3* have been seen in cancers, however our data do not suggest that pLoF patients are at risk for cancer; there are not enough data to suggest an association.

There are limitations to our attempt to aggregate a consensus phenotype for this variant type that are not unique among initial descriptions of novel syndromes, including: a moderate number of cases, variable depth of clinical data, the potential for ascertainment bias, and a lack of clinical histories across the full lifespan. Further clarity will be realized with additional cases over time, buoyed by even larger-scale genomic data from emerging cohorts numbering in the millions of participants (e.g. The All of Us Research Program Investigators^72^).

Finally, our work suggests the existence of additional considerably haploinsufficient genes, LoF of which yield yet-undiscovered mild-moderate or nonspecific phenotypes that will be ascertained only by careful, hybrid studies marrying genomic and patient-level data.

## Data Availability

All data produced in the present work are contained in the manuscript.

## Acknowledgements

The authors thank: the patients and families who participated in this study; Dr. Jason Flannick, Emma Sherrill, Minh Nguyen, Aubrie Soucy, and the Cornelia de Lange Syndrome Foundation, Inc. for general support of the project; and Drs. Jennifer Gerton, Zuzana Tothova, and Kaitlin Samocha for helpful discussion. P.M.B. is supported by award K08NS117891 from the US National Institute of Neurological Disorders and Stroke (NINDS), and an award from the Boston Children’s Hospital Office of Faculty Development.

K.N.W.F. and the Clinic for Cornelia de Lange Syndrome and Related Disorders are supported by gifts from Peter and Kathy Wagner and Julie and Frank Mairano. M.E.T, P.M.B., L.W., and D.B-I were supported by grants from the US National Institutes of Health (NIH) (U01HG011755, R01MH115957, HD081256, and P50HD104224). D.B is supported by T32HG002295 from the US National Human Genome Research Institute. S.S. is supported by award K23NS119666 from the US NINDS. T.B.H. is supported by the Deutsche Forschungsgemeinschaft (German Research Foundation) – 418081722, 433158657. Some sequencing and analysis was provided by the Broad Institute Center for Mendelian Genomics and was funded by NIH grants UM1HG008900 and R01HG009141. K.M.B is supported by US National Eye Institute (R01EY026904, R01EY012910, P30EY014104) and the Foundation Fighting Blindness (EGI-GE-1218-0753-UCSD). This work was supported in part by the National Institutes of Health of the United States of America grants R01HD078641 and P50HD096723 to D.F.C. S.B. is supported by the NIHR Manchester Biomedical Research Centre (NIHR203308). A.S. is supported by RC2DK122533. M.S-B was supported by NHLBI R01HL143295. Analysis by E.G., S.T. and A.O’D.L was supported by U01HG0011755 and R01HG009141. This work was supported by the Boston Children’s Hospital Rare Disease Cohorts Initiative (CRDC). This study makes use of data generated by the DECIPHER community. A full list of centers who contributed to the generation of the data is available from https://deciphergenomics.org/about/stats and via email from. Funding for the DECIPHER project was provided by Wellcome (grant number WT223718/Z/21/Z).

## Data and Code Availability

The published article and its supplement list all datasets analyzed during this study.

