## Supplemental Tables and Figures for "Heterozygous loss-of-function *SMC3* variants are associated with variable and incompletely penetrant growth and developmental features"

### **Table S1. Variant details of heterozygous *SMC3* predicted LoF variants in cases.**

**Table S2. Heterozygous *SMC3* pLoF variants among genomic cohorts.**

**Table S3. gnomAD and UKBB pLoF variants.**

**Table S4. *SMC3* missense variants among mostly male infertility cases.**

**Table S5. Key gene lists for expression analyses.**

**Table S6. *SMC3*, *SMC1A*, *RAD21* Perturb-seq parameters.**

### **Supplemental Figures**

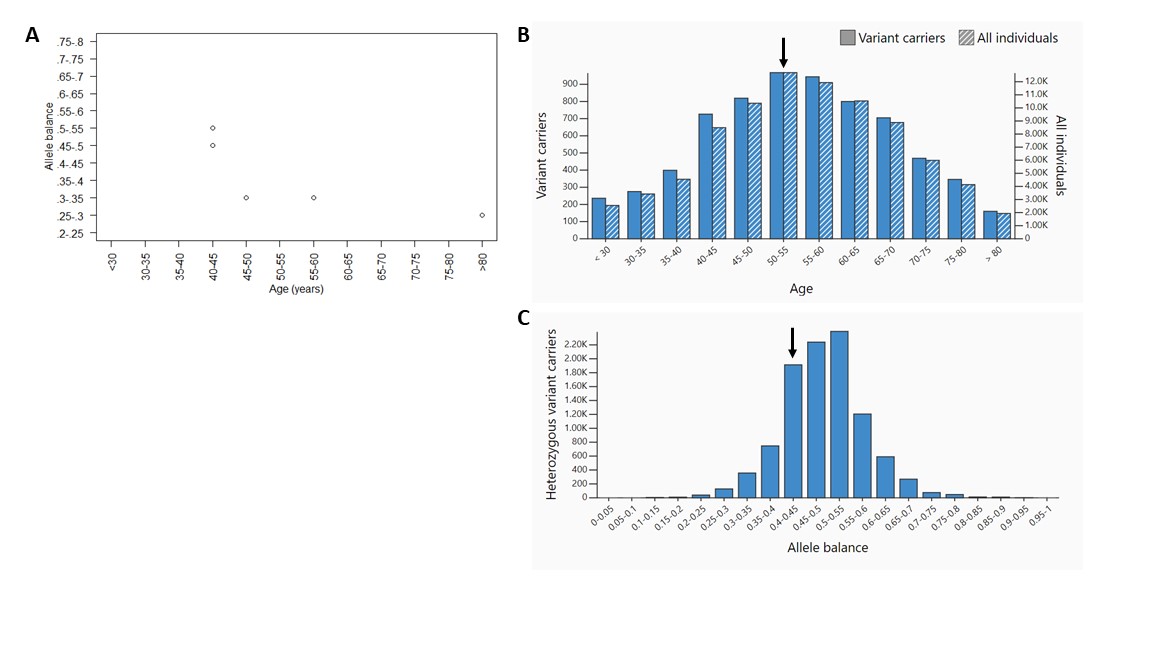

**Figure S1. *SMC3* pLoF variants identified via gnomAD do not exhibit signs of clonal hematopoiesis. a.** Both age and allele balance were available for 5/9 pLoF *SMC3* variants in gnomAD passing quality filters.^1^ None of these variants overlap with *SMC3P1.* **b.** Age distribution of heterozygous carriers of an *SMC3* synonymous, ClinVar “benign” control variant, p.(Tyr455Tyr) (hg19 10-112349422-T-C, hg38 10-110589664-T-C) with an allele frequency of 0.0442 in gnomAD, compared with all individuals. Only data from exome sequencing are shown. The arrow demonstrates the mean age category of five *SMC3* pLoF variant carriers, in which >80y was given equal weight to other age bins. **c.** Allele balance for heterozygous carriers of the p.(Tyr455Tyr) variant. Only data from exome sequencing are shown. The arrow demonstrates the mean allele balance range for nine *SMC3* pLoF variants.

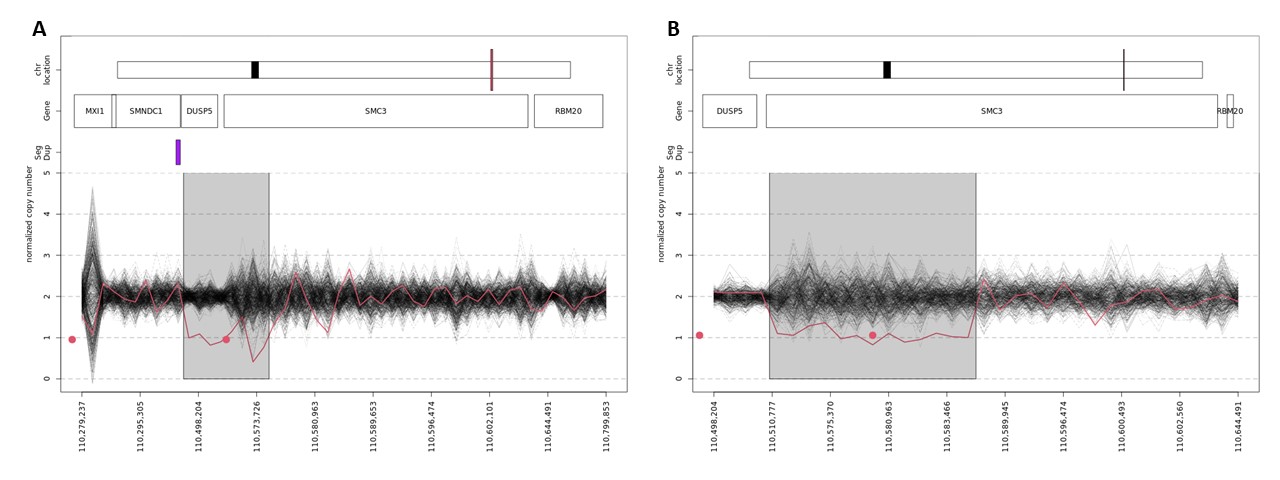

**Figure S2. Deletions involving *SMC3* exons among individuals in the UKBB.** Two deletions were discovered among 196,869 individuals passing quality filters. The red lines show the copy numbers inferred by GATK-gCNV. In each panel, the gray rectangle depicts the boundaries of the deletion call, and the red dots display the mean copy number across the deleted segment. The black lines show the copy numbers inferred by GATK-gCNV for the other samples processed in the same batch as the depicted deletion. The horizontal axis displays the genomic positions of the inferred copy number calls, which have been rescaled so the captured exome sequencing intervals are evenly spaced across the plotted region. The top of each panel shows the relative positions of the plotted region (tall, thin rectangle) and centromere (short, thick rectangle) on the relevant chromosome as well as the locations of genes and segmental duplications in the plotted region.

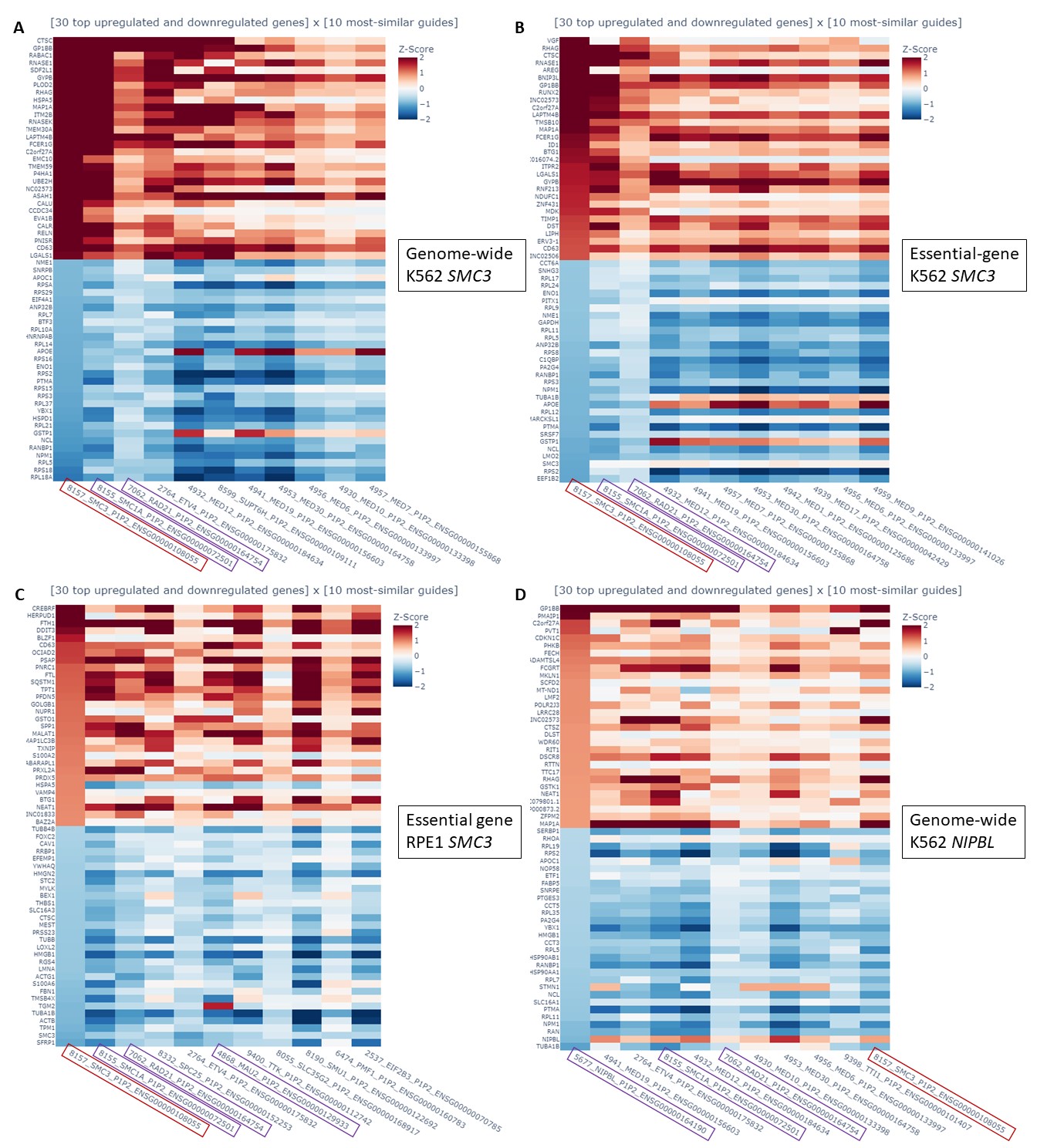

### **Figure S3. The gene expression profile of *SMC3* knockdown is correlated with that of other cohesin genes across experiments and cell types in Perturb-seq data.** Perturb-seq data were derived from Replogle et al.^2^ and plotted via (<https://gwps.wi.mit.edu>). The top two matches to *SMC3* knockdown (red) in each experiment are the other cohesin ring components *RAD21* and *SMC1A*, and there are similarities with the cohesin loaders *MAU2* and *NIPBL* as well (these other cohesin genes in purple).

### **References (Supplement)**

1	Karczewski, K. J. *et al.* The mutational constraint spectrum quantified from variation in 141,456 humans. *Nature* **581**, 434-443 (2020). <https://doi.org:10.1038/s41586-020-2308-7>

2	Replogle, J. M. *et al.* Mapping information-rich genotype-phenotype landscapes with genome-scale Perturb-seq. *Cell* **185**, 2559-2575 e2528 (2022). <https://doi.org:10.1016/j.cell.2022.05.013>
